# Effects of a spore-forming probiotic blend on bowel habits and physical well-being in adults with functional constipation: a randomized, double-blind, placebo-controlled trial

**DOI:** 10.1101/2025.11.05.25339581

**Authors:** Hyung Gyu Park, Han Bin Lee, Haeseong Park, Minji Kang, Minkyung Bok, Yeongtaek Hwang, Kyuho Jeong, Sungho Maeng, Hyunjung Lim, Jin Seok Moon

## Abstract

We aimed to evaluate the efficacy and safety of a spore-forming probiotic blend containing *Clostridium butyricum* IDCC 1301, *Weizmannia coagulans* IDCC 1201, and *Bacillus subtilis* IDCC 1101 for improving bowel function and well-being in adults with functional constipation (FC). In a randomized, double-blind, placebo-controlled trial, 78 adults with FC (Rome IV criteria) received either probiotic blend (n = 40) or placebo (n = 38) daily for 4 weeks. Primary outcomes were changes in weekly spontaneous bowel movements (WSBM) and stool form. Secondary outcomes included physical functioning scores from the 36-Item Short Form Health Survey. The probiotic blend group showed significant improvements in irritant bowel movements (p = 0.0458), incomplete evacuation (p = 0.0374), and abdominal pain before defecation (p = 0.0090). Stool consistency shifted toward normal types (Bristol types 3–4, p = 0.0176). Physical functioning improved only in the probiotic blend group (p = 0.0300). Probiotic blend effectively alleviated symptoms of FC and improved physical well-being.

## Introduction

Functional constipation (FC) is a common gastrointestinal disorder diagnosed based on the Rome IV criteria [1]. FC affects approximately 14% of the global population and is characterized by abdominal discomfort or pain, distension, and a sensation of incomplete evacuation [2–4]. The pathogenesis of FC is multifactorial, involving genetic predisposition, lifestyle habits, and psychological factors [5]. Recently, alterations in gut microbiota composition have emerged as potential risk factors for the development of FC [6]. Several studies have reported that gut microbiota dysbiosis—particularly a reduced abundance of *Bifidobacteria*, *Lactobacillus*, *Bacteroides*, and *Prevotella*—is associated with FC [7]. Accordingly, the underlying mechanisms of FC are complex and not yet fully understood.

Despite the availability of diverse therapeutic strategies, the management of FC remains challenging [8]. Conventional pharmacological treatments often fail to address the multifactorial causes of FC and may lead to inconsistent symptom responses among individuals [8]. Emerging evidence sup-ports the central role of gut microbiota in gastrointestinal homeostasis and highlights microbial balance as a critical factor in the effective management of FC [9]. In this context, probiotic intake has gained considerable attention as a promising adjunctive therapy. By modulating gut microbiota com-position and enhancing mucosal barrier function, probiotics may offer a non-pharmacological approach to alleviate symptoms and improve bowel function in individuals with FC [10].

BIOVITA^®^ Blend is the first probiotic medicinal product developed in Korea in 1959 [11]. The probiotic blend consists of *Bacillus subtilis* IDCC 1101 (*B. subtilis* IDCC 1101), *Weizmannia coagulans* IDCC 1201 (*W. coagulans* IDCC 1201), and *Clostridium butyricum* IDCC 1301 (*C. butyricum* IDCC 1301). Among them, *B. subtilis* IDCC 1101 is a promising probiotic strain because of its high stability and demonstrated safety [12]. *W. coagulans* IDCC 1201 was also evaluated for safety and showed antibacterial activity and anti-inflammatory effects [13,14].

*C. butyricum* IDCC 1301 is a representative butyric acid-producing bacterium [15]. In particular, butyric acid produced by *C. butyricum* serves as a key energy source for colonic epithelial cells, helping maintain intestinal health by increasing mucus secretion and enhancing barrier function [3]. Considering that these three strains are spore-forming and colonize different regions of the intestine, their combination provides comprehensive benefits throughout the intestinal tract, from the small to the large intestine [11].

Previous studies revealed that probiotic blend alleviates loperamide-induced constipation [16,17]. Oral administration of probiotic blend increased the fecal pellet count, fecal water content, and gastrointestinal transit ratio in Sprague-Dawley rats [16,17]. While numerous preclinical studies demonstrate the usefulness of each strain and the efficacy of probiotic blend against FC, clinical validation through human studies remains insufficient.

In this study, we investigated the effects of a spore-forming probiotic blend on improving bowel habits in participants diagnosed with FC (Roma IV criteria). Efficacy—including defecation activity and quality of life (QoL)—and safety were evaluated as outcome variables.

## Materials and Methods

### Study design

This randomized, double-blind, placebo-controlled trial conducted from September 2023 to March 2024 received approval from the Institutional Review Board of Kyung Hee University Korean Medicine Hospital (KOMCIRB 2023-05-001) and was registered with the Clinical Research Information Service (https://cris.nih.go.kr/cris/index/index.do) under registration number KCT0010085.

Participants who voluntarily provided written informed consent were enrolled after passing the screening test and meeting the eligibility criteria. At visit 1 (week 0), eligible participants were randomly allocated to the intervention or control group. Random allocation was performed in a 1:1 ratio between the intervention and control groups. Participants consumed either the probiotic blend or placebo once daily for 4 weeks while maintaining their usual lifestyle. Over the 4-week study period, participants visited Kyung Hee University Korean Medicine Hospital three times (week 0, week 2, and week 4) for efficacy and safety assessments.

### Participants

Participants were recruited through hospital bulletin board postings and external advertisements. Individuals visited the hospital for eligibility assessment based on the predefined inclusion and exclusion criteria. All participants were provided with a detailed explanation of the study protocol, and written consent was obtained voluntarily before enrolment.

The inclusion criteria were as follows: (1) age 19–65 years; (2) diagnosed with FC according to the Rome IV criteria; (3) voluntarily agreed to participate in the study and signed the informed consent form.

The exclusion criteria were as follows: (1) diagnosed with irritable bowel syndrome (IBS); (2) consumption of medications, herbal remedies, or probiotic supplements known to influence constipation symptoms (e.g., bulk-forming laxatives, stool softeners, rhubarb, aloe, probiotics, etc.) within the past 2 weeks; (3) diagnosed with or currently receiving treatment for acute or chronic gastrointestinal diseases, including Crohn’s disease, celiac disease, ulcerative colitis, or colorectal malignancies; (4) history of gastrointestinal or hepatobiliary surgery, except for appendectomy; (5) history of viral hepatitis, malignant tumors, mental disorders, or autoimmune diseases; (6) following a dietary regimen for weight control; (7) patients with diabetes whose fasting blood glucose remains ≥ 126 mg/dL despite medication use; (8) patients with hypertension whose systolic blood pressure remains ≥ 160 mmHg or diastolic blood pressure remains ≥ 100 mmHg despite antihypertensive medication; (9) aspartate aminotransferase, alanine aminotransferase, or serum creatinine levels exceeding twice the normal reference range; (10) thyroid dysfunction; (11) pregnant or lactating women, or those planning to conceive within 6 months; (12) hypersensitivity or allergies to the test product or any of its ingredients; (13) participation in another clinical trial or human study within the past month; (14) limited reading ability; (15) deemed unsuitable for participation at the discretion of the principal investigator.

### Sample size calculation

The sample size for this study was determined based on the results of a previous study conducted in patients with functional constipation. In that study, the weekly bowel movement frequency after 4 weeks of intervention was significantly higher in the intervention group (5.28 ± 1.93 times/week) compared with the control group (3.89 ± 1.79 times/week) (p < 0.05) [18]. Using the G*Power 3.1.9.7 (Heinrich Heine University Düsseldorf, Düsseldorf, Germany), the minimum required sample size was calculated to be 60 participants, based on an effect size of 0.75, a significance level of 0.05, and a statistical power of 0.80. To account for an anticipated dropout rate of 25%, the final sample size was set at 80 participants, with 40 allocated to each group.

### Randomization

Participants were randomly assigned to the intervention and control groups in a 1:1 ratio using a probability-based method. An independent statistician generated randomization numbers, employing a random number generator to implement block randomization. Participants were assigned a Randomization Number based on the randomization table in the order of enrolment and subsequently allocated to either the intervention or control group. Study products were distributed according to the assigned codes generated through randomization.

### Preparation of investigation product

For the seed culture, 1% (v/v) of glycerol stock of *C. butyricum* IDCC 1301, *W. coagulans* IDCC 1201, and *B. subtilis* IDCC 1101 was suspended and incubated in De Man, Rogosa, and Sharpe broth at 45 °C in a 50 L fermenter for 12 h. Ten per cent (v/v) of the seed culture was transferred into a commercial medium and incubated in a 500 L fermenter at 30 rpm for 6 h. The main culture was performed with 2% of preculture under the conditions mentioned for 16 h in a 20 kL fermenter (Fig 1). The biomass and culture media were separated by continuous centrifugation at a constant rate of 10 kL/h and 8,000 rpm, yielding 100–150 kg of *C. butyricum* IDCC 1301, *W. co-agulans* IDCC 1201, and *B. subtilis* IDCC 1101. The cell concentrations were freeze-dried at a temperature range of -80 to -25 °C under a pressure of less than -300 mmHg. Finally, the freeze-dried microbial cake was finely ground and sieved to obtain particles < 50 μm. *C. butyricum* IDCC 1301, *W. coagulans* IDCC 1201, and *B. subtilis* IDCC 1101 were mixed in specific proportions and the particles were then encapsulated. The compositions of the probiotic blend and placebo products are listed in S1 Table. The investigation product contained more than 1.0 × 109 colony-forming units of probiotic blend complex per capsule. probiotic blend and placebo capsules were prepared in similar sizes and colors. Participants were instructed to take one capsule per day for 4 weeks.

**Fig 1.**
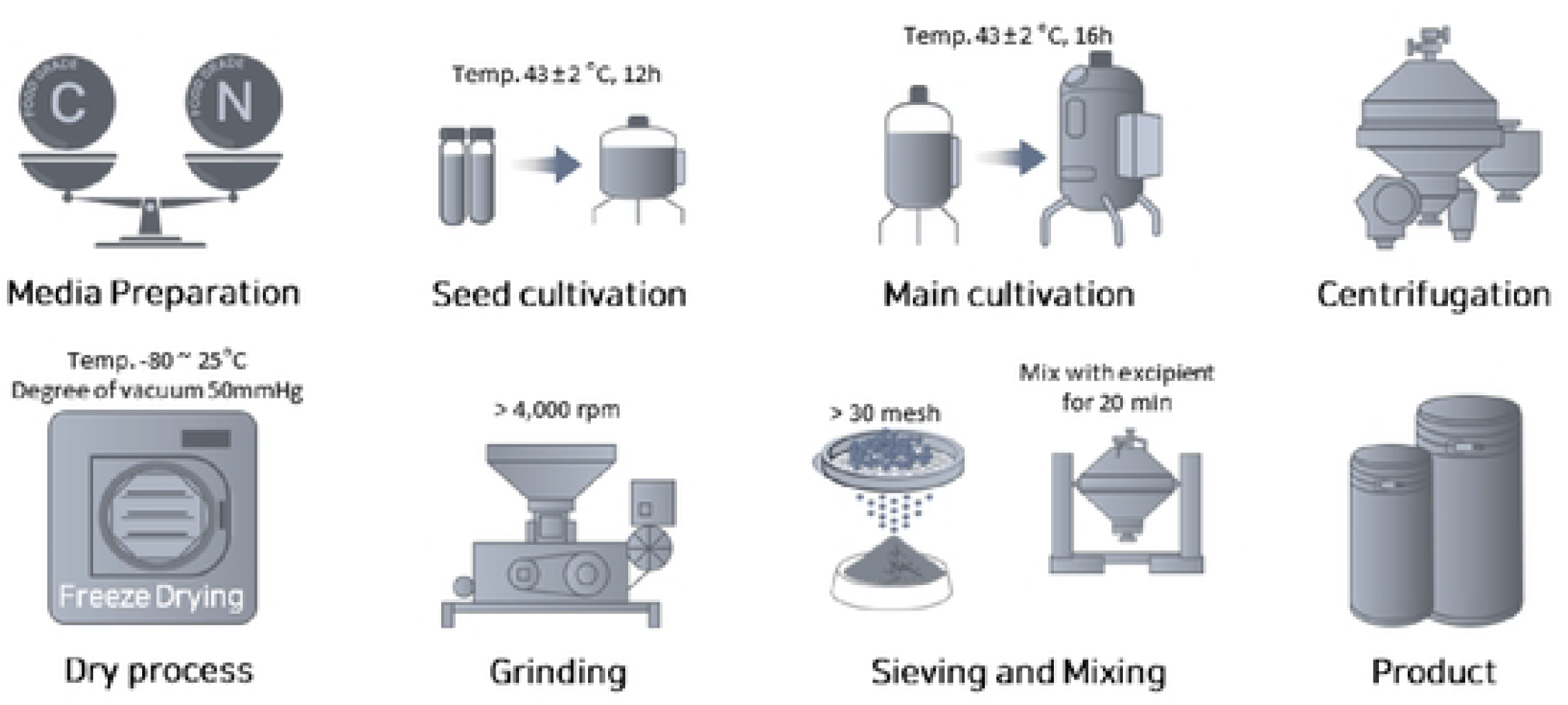
The Manufacturing Process for probiotic blend Products. C, carbon source. N, nitrogen source.

### Outcome

Bowel habits were assessed using a bowel activity questionnaire at Visit 1 (week 0), Visit 2 (week 2), and Visit 3 (week 4). The questionnaire items were designed with reference to the guidelines pro-vided by the Ministry of Food and Drug Safety, incorporating relevant questions and surveys on bowel habits [19]. The assessment included weekly bowel movement frequency, stool volume, abdominal pain, sensation of incomplete evacuation, bloating during defecation, abdominal discomfort after bowel movements, and discomfort due to constipation. These parameters were measured using a Visual Analog Scale ranging from 0–10. Stool consistency (Shape) was recorded based on the Bristol Stool Scale [20]. Bowel movement frequency was defined as the number of spontaneous bowel movements occurring in the previous week, excluding any bowel movements induced by enemas, suppositories, or digital maneuvers. To minimize external influences, participants were instructed to maintain their usual dietary and exercise habits throughout the study.

QoL was evaluated using the 36-item Short Form Health Survey (SF-36), a standardized questionnaire for assessing individual QoL [21]. The SF-36 consists of 36 items covering eight domains: physical functioning, role limitations due to physical health, bodily pain, general health perception, vitality, social functioning, role limitations due to emotional health, and mental health. Participants completed the SF-36 questionnaire at visit 1 (week 0), visit 2 (week 2), and visit 3 (week 4). The responses were then coded and scored to evaluate each domain.

### Statistical analysis

The primary efficacy analysis was conducted using the per-protocol (PP) set. Continuous variables were reported as mean ± standard deviation (SD), whereas categorical variables were presented as numbers (%). Categorical variables were analyzed using the chi-squared test or Fisher’s exact test, and continuous variables were analyzed using the Mann-Whitney U test. The normality of efficacy data distribution was assessed using the Shapiro-Wilk test. Differences in changes between the intervention and control groups were analyzed using the Mann-Whitney U test for continuous variables. Within-group changes between week 0 and week 4 were assessed using Wilcoxon’s signed-rank test for continuous variables and McNemar’s test for categorical variables. Analysis of covariance (AN-COVA) was conducted to adjust for participant characteristics or baseline values in efficacy assessments. Statistical significance was defined as a p-value < 0.05. All statistical analyses were performed using SAS (Version 9.4, SAS Institute, Cary, North Carolina, USA).

## Results

### Study population and baseline characteristics

A total of 80 participants were enrolled and randomly assigned to either the probiotic blend group (n = 40) or the placebo group (n = 40). However, two participants withdrew from the placebo group, resulting in a final sample of 40 participants in the probiotic blend group and 38 in the placebo group for the per-protocol analysis (Fig 2). Participant recruitment occurred from September 2023 to March 2024, and all follow-up assessments were completed by April 2024. The trial was completed as planned, with no early termination or safety concerns. Both the probiotic blend and placebo capsules were identical in size, color, and packaging, ensuring complete blinding throughout the study. Participants were instructed to take one capsule daily for 4 weeks while maintaining their usual diet and lifestyle. Adherence was monitored through capsule counts and participant diaries. The mean compliance rate was 98.0% in the probiotic blend group and 97.6% in the placebo group, indicating high fidelity to the intervention protocol. Baseline characteristics were generally well balanced between the groups regarding age, gender, occupation, lifestyle factors (including alcohol consumption, smoking, and exercise), and dietary habits (such as frequency of meals, snack intake, and water consumption). Energy intake and physical activity levels showed no significant differences between the probiotic blend and placebo groups during the study period (S2 Table and S3 Table). However, significant differences were observed in disease history (probiotic blend group: 25.0% vs. placebo group: 2.6%, p = 0.0046) and sleep duration (a higher proportion of participants in the probiotic blend group reported ≤ 7 h of sleep, p = 0.0197). Given these baseline discrepancies, ANCOVA was performed to adjust for potential confounding effects in the primary outcome analysis (Table 1).

**Fig 2.**
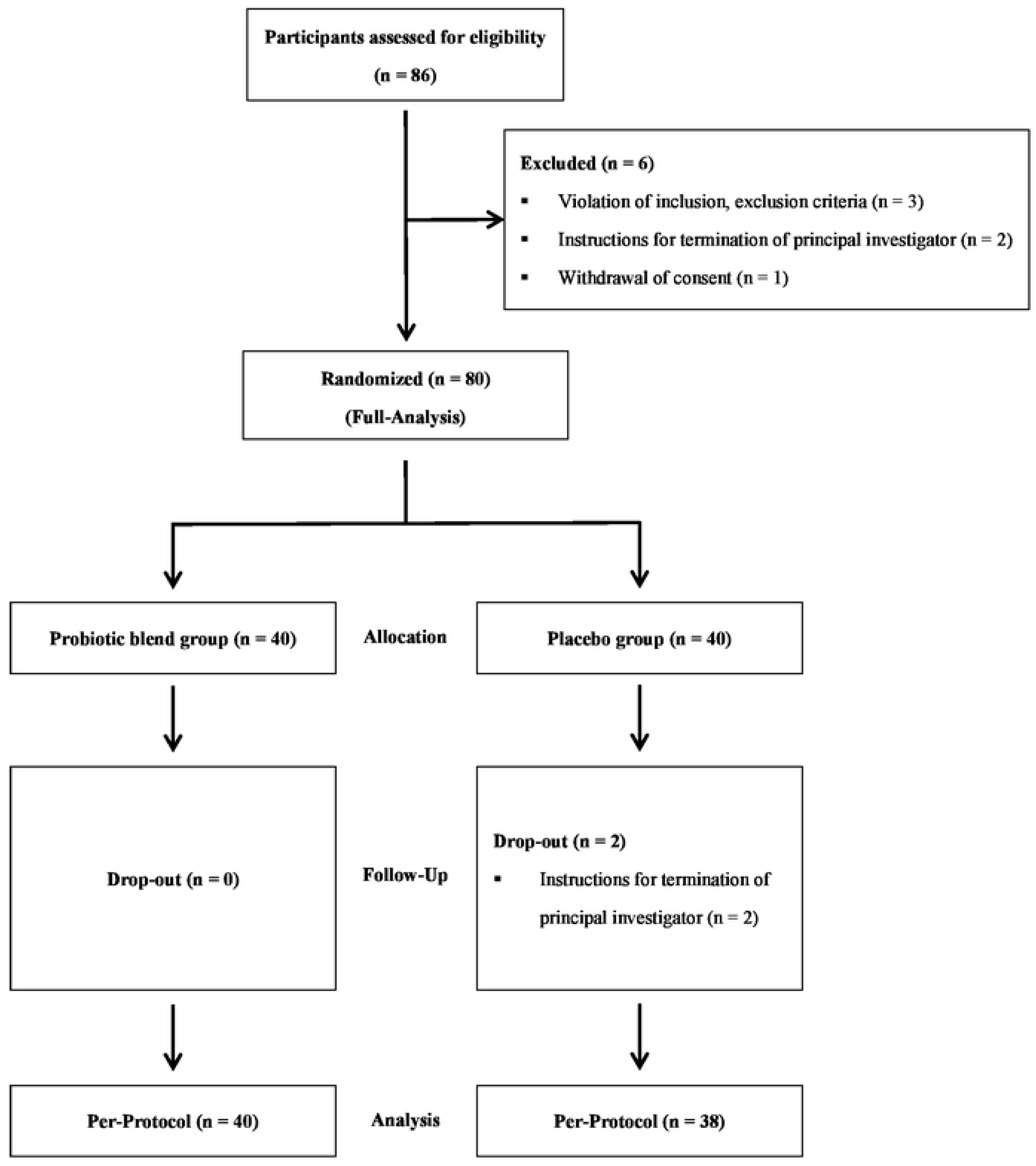
Flow of Study.

**Table 1.**
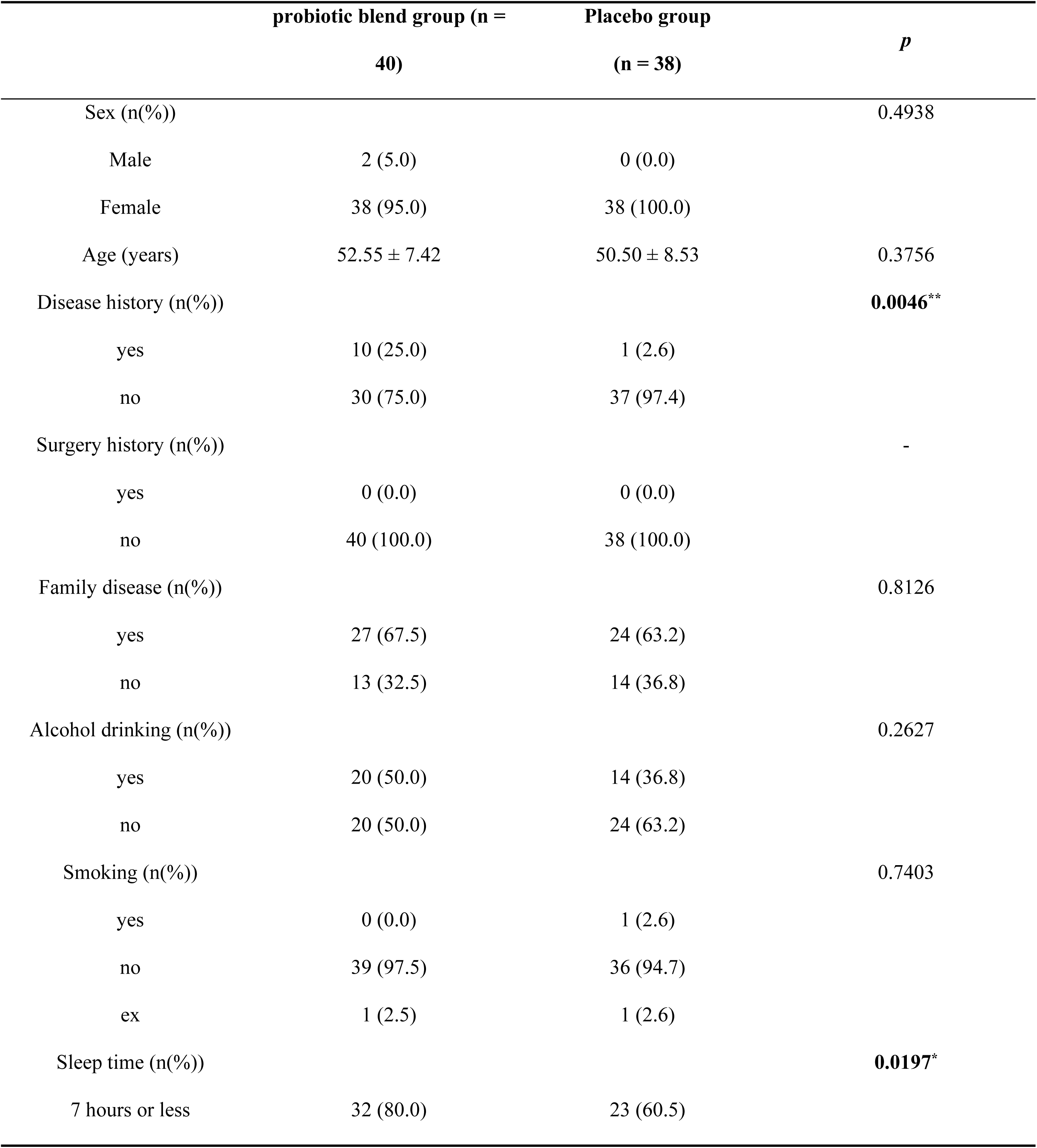
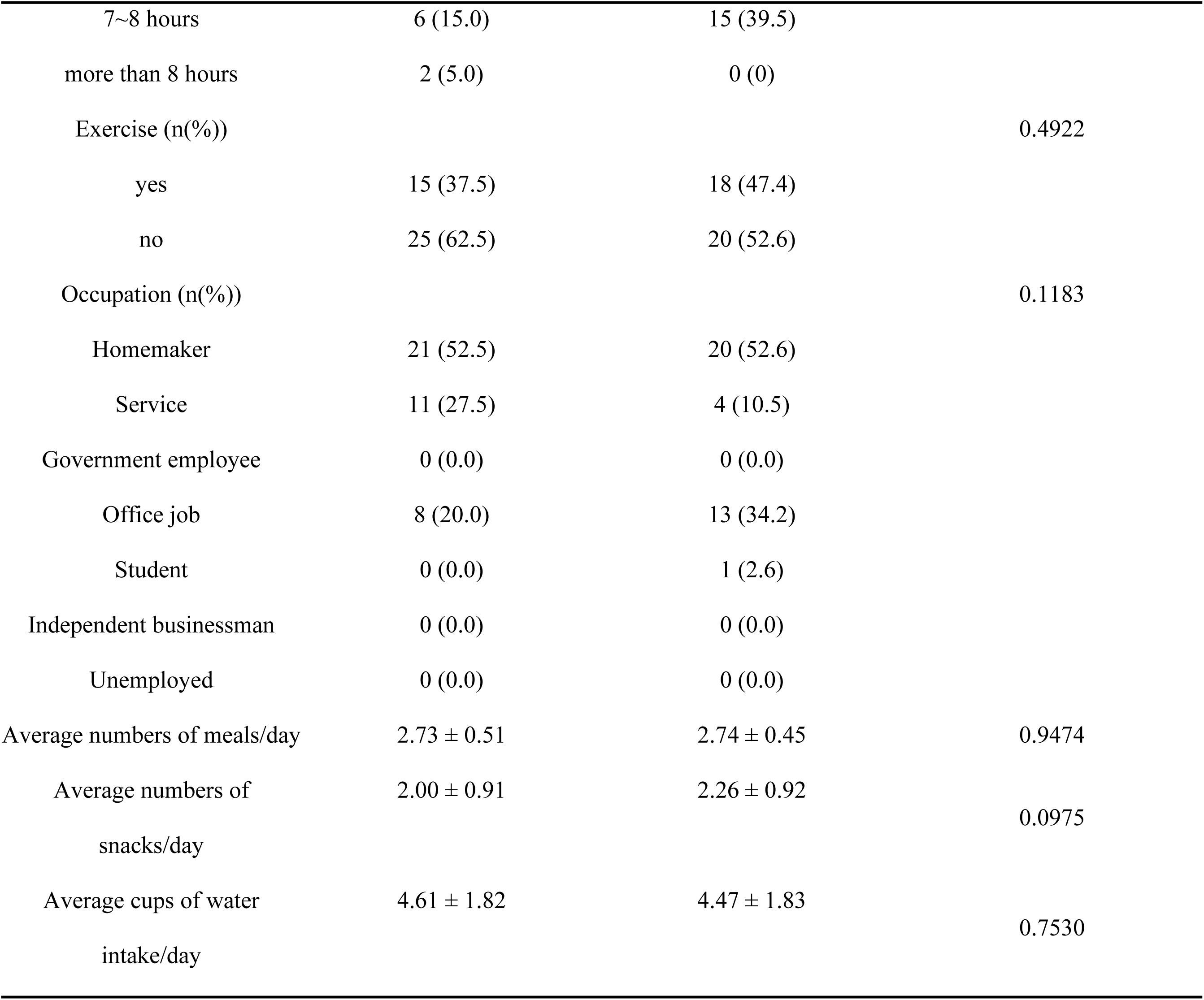
General Characteristics at Baseline.

### Changes in bowel function

The analysis of bowel function revealed that probiotic blend supplementation was associated with significant improvements in defecation-related discomfort. As shown in Fig 3, the frequency of irritant bowel movements was significantly reduced in the probiotic blend group compared with the placebo group (-2.63 ± 2.13 vs. -1.89 ± 1.98, p = 0.0458). Likewise, the frequency of incomplete bowel movements showed a greater reduction in the probiotic blend group (-2.75 ± 2.23 vs. -1.74 ± 2.18, p = 0.0374), indicating improved defecation satisfaction. Participants in the probiotic blend group also experienced a significant reduction in the frequency of abdominal pain before bowel movements (-1.13 ± 1.54), whereas no notable change was observed in the placebo group (-0.11 ± 1.71, p = 0.0090).

**Fig 3.**
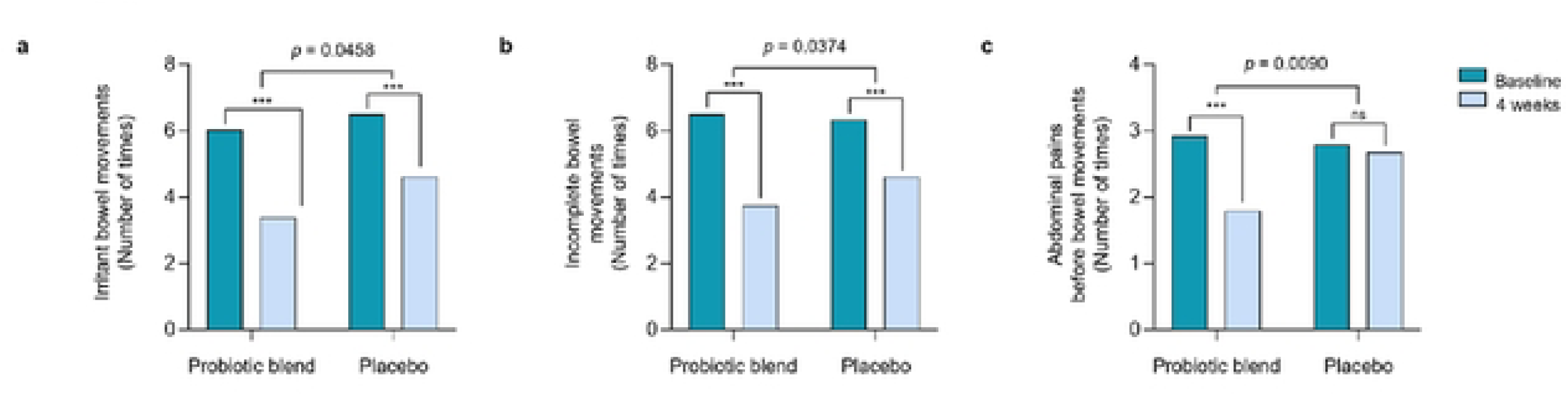
Changes in Bowel Function from Baseline to Week 4. The parameters include: (a) number of times of irritant bowel movements, (b) number of times when bowel movements felt incomplete, and (c) number of times of abdominal pain before bowel movements. Data are expressed as mean values. Significant difference between baseline and 4 weeks data by Wilcoxon signed-rank test at * < 0.05, ** < 0.01, *** < 0.001. P-values were calculated using Mann–Whitney U test. The difference in changes between groups was calculated after adjustment for disease history and sleep time using analysis of covariance. ns, not significant

All parameters related to bowel function are summarized in S4 Table. Although weekly bowel movement frequency increased in both groups, no statistically significant difference was observed between the probiotic blend and placebo groups (1.00 ± 0.99 vs. 0.95 ± 0.96, p = 0.6532).

A significant reduction in the severity of abdominal pain was observed in the probiotic blend group, whereas no such change was seen in the placebo group. However, the between-group difference did not reach statistical significance (2.38 ± 1.92 vs. 3.32 ± 2.27, p = 0.0778). Other parameters, including the frequency of abdominal pain during bowel movements, gas volume, discomfort after bowel movements, and discomfort caused by constipation, exhibited numerical improvements in both groups, but these changes were not statistically significant.

### Changes in stool characteristics and defecation patterns

In addition to improvements in bowel function, probiotic blend supplementation significantly influenced stool consistency. At baseline, stool form, classified according to the Bristol Stool Scale, was comparable between the two groups. However, by the end of the study, the probiotic blend group exhibited a significant shift toward a more optimal stool consistency (p = 0.0176). A higher proportion of the probiotic blend group participants had normal stool shape (shapes 3–4), whereas changes in the placebo group were less pronounced. No significant difference was found in defecation time (p = 0.0556), although a trend toward shorter durations in the probiotic blend group was observed. Similarly, the amount of stool remained comparable between groups (p = 0.7787), suggesting that probiotic blend primarily enhances stool quality rather than total stool output (Table 2).

**Table 2.**
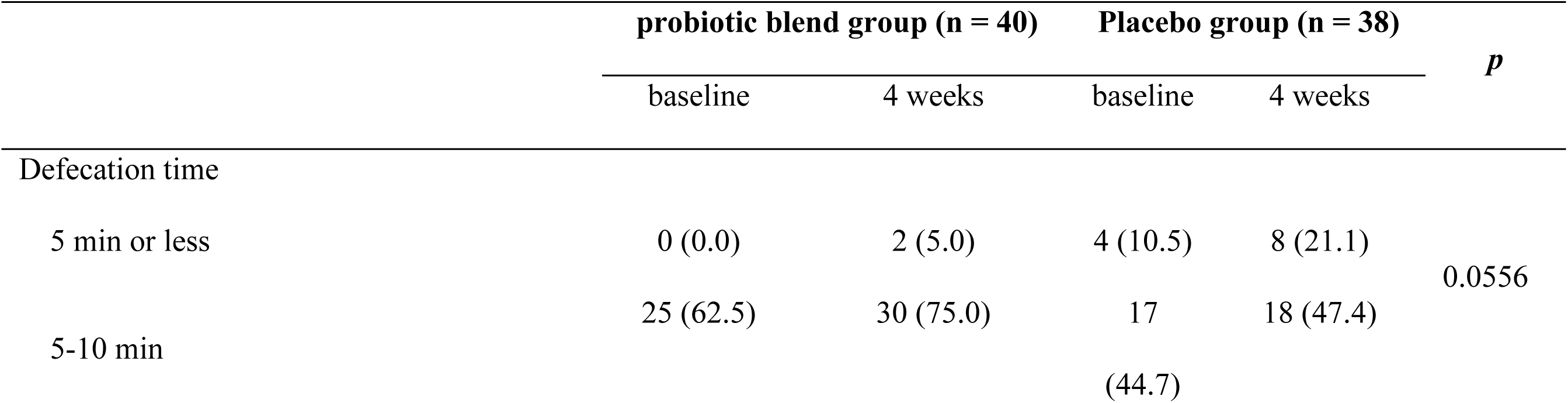
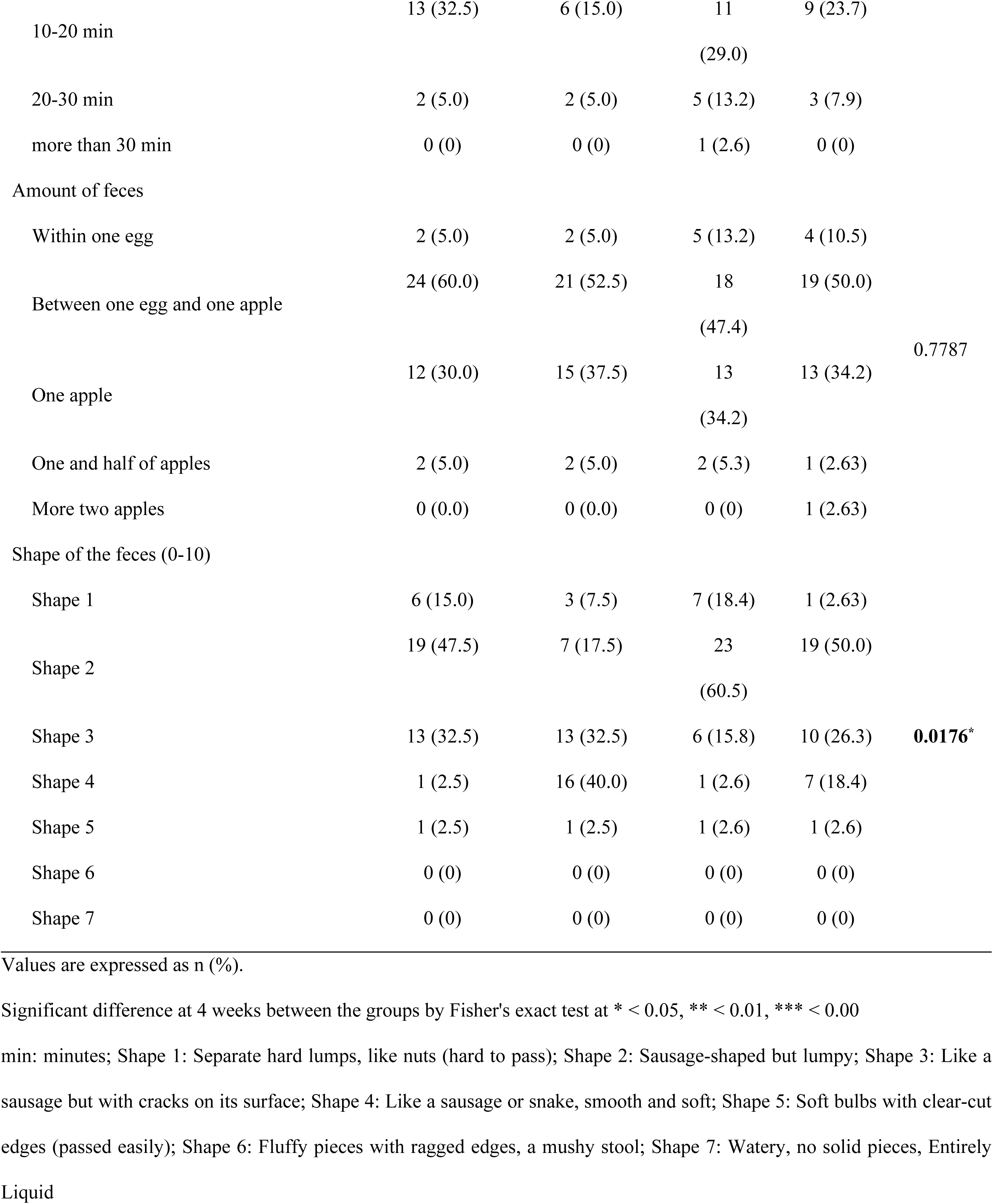
Changes in Stool Characteristics and Defecation Patterns.

|  | probiotic blend group (n = 40) |  | Placebo group (n = 38) |  | *p* |
| --- | --- | --- | --- | --- | --- |
|  | baseline | 4 weeks | baseline | 4 weeks |  |
| Defecation time |  |  |  |  |  |
| 5 min or less | 0 (0.0) | 2 (5.0) | 4 (10.5) | 8 (21.1) | 0.0556 |
| 5-10 min | 25 (62.5) | 30 (75.0) | 17 (44.7) | 18 (47.4) |  |
| 10-20 min | 13 (32.5) | 6 (15.0) | 11<br>(29.0) | 9 (23.7) |  |
| 20-30 min | 2 (5.0) | 2 (5.0) | 5 (13.2) | 3 (7.9) |  |
| more than 30 min | 0 (0) | 0 (0) | 1 (2.6) | 0 (0) |  |
| Amount of feces |  |  |  |  |  |
| Within one egg | 2 (5.0) | 2 (5.0) | 5 (13.2) | 4 (10.5) |  |
| Between one egg and one apple | 24 (60.0) | 21 (52.5) | 18<br>(47.4) | 19 (50.0) | 0.7787 |
| One apple | 12 (30.0) | 15 (37.5) | 13<br>(34.2) | 13 (34.2) |  |
| One and half of apples | 2 (5.0) | 2 (5.0) | 2 (5.3) | 1 (2.63) |  |
| More two apples | 0 (0.0) | 0 (0.0) | 0 (0) | 1 (2.63) |  |
| Shape of the feces (0-10) |  |  |  |  |  |
| Shape 1 | 6 (15.0) | 3 (7.5) | 7 (18.4) | 1 (2.63) |  |
| Shape 2 | 19 (47.5) | 7 (17.5) | 23<br>(60.5) | 19 (50.0) |  |
| Shape 3 | 13 (32.5) | 13 (32.5) | 6 (15.8) | 10 (26.3) | **0.0176\*** |
| Shape 4 | 1 (2.5) | 16 (40.0) | 1 (2.6) | 7 (18.4) |  |
| Shape 5 | 1 (2.5) | 1 (2.5) | 1 (2.6) | 1 (2.6) |  |
| Shape 6 | 0 (0) | 0 (0) | 0 (0) | 0 (0) |  |
| Shape 7 | 0 (0) | 0 (0) | 0 (0) | 0 (0) |  |

Values are expressed as n (%).
Significant difference at 4 weeks between the groups by Fisher's exact test at \* < 0.05, \*\* < 0.01, \*\*\* < 0.00
min: minutes; Shape 1: Separate hard lumps, like nuts (hard to pass); Shape 2: Sausage-shaped but lumpy; Shape 3: Like a sausage but with cracks on its surface; Shape 4: Like a sausage or snake, smooth and soft; Shape 5: Soft bulbs with clear-cut edges (passed easily); Shape 6: Fluffy pieces with ragged edges, a mushy stool; Shape 7: Watery, no solid pieces, Entirely Liquid

### Changes in QoL

The effects of probiotic blend supplementation extended beyond bowel function and stool characteristics, as evidenced by improvements in QoL assessments. A significant improvement was observed in physical functioning scores, which increased in the probiotic blend group (1.38 ± 14.32) but declined in the placebo group (-3.16 ± 13.73, p = 0.0300). However, no significant differences were found between groups in social functioning (p = 0.2412), mental health, or role-emotional scores, indicating that the primary benefits of probiotic blend may be physiological rather than psychological (Table 3).

**Table 3.**
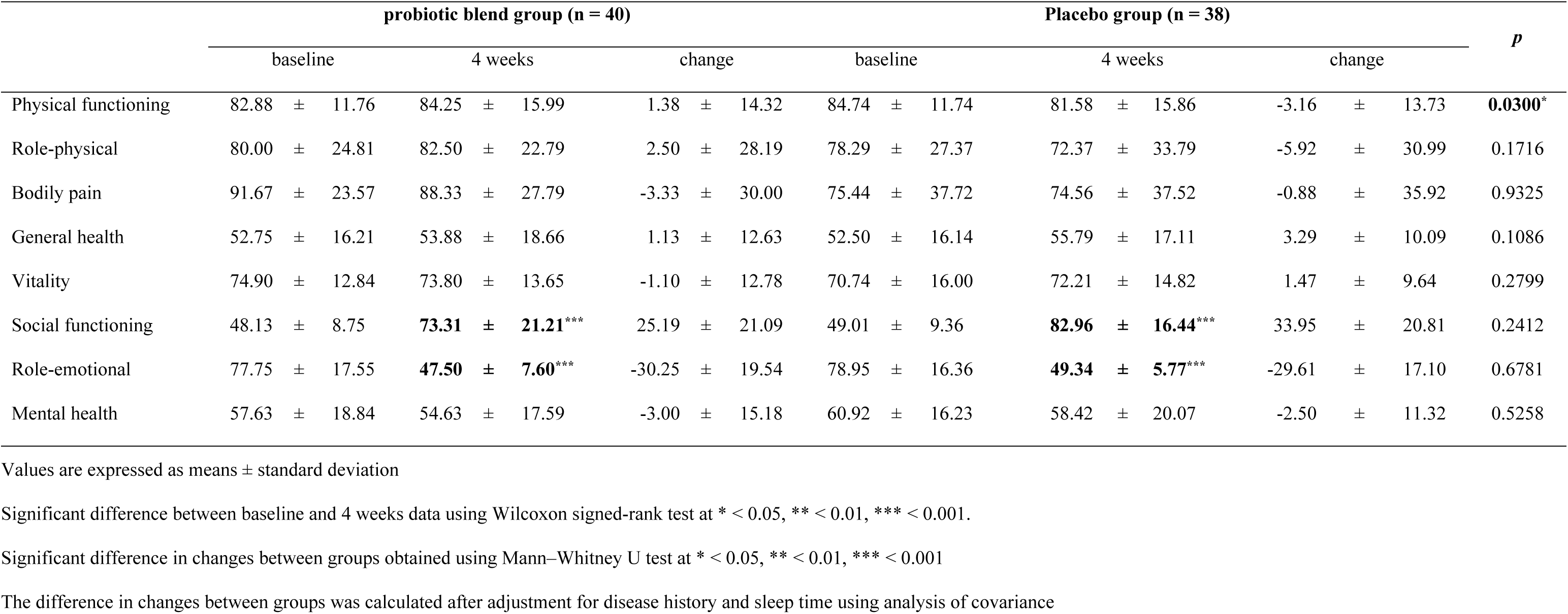
Changes in Quality of Life.

## Discussion

In this randomized, double-blind, placebo-controlled clinical trial, we assessed whether a four-week oral intake of a multi-strain, spore-forming probiotic blend could ease bowel-related discomfort in healthy adults experiencing mild abdominal symptoms. A total of 80 participants were enrolled, with 78 completing the study. Demographic characteristics such as age and sex were generally comparable between the two groups (Table 1). However, differences in sleep duration and medical history were observed and statistically adjusted using ANCOVA.

As illustrated in Fig 3, participants receiving the probiotic blend experienced notable reductions in defecation difficulty, feelings of incomplete evacuation, and pre-defecation abdominal pain. These outcomes suggest improved bowel comfort. Stool consistency also improved (Table 2), indicating that the probiotic may help regulate stool formation— something particularly relevant for individuals with irregular bowel habits. Improvements weren’t limited to digestive symptoms alone. Physical functioning scores also increased in the probiotic group (Table 3), pointing to potential benefits for overall quality of life, especially considering how bowel discomfort often restricts physical activity.

Interestingly, while overall bowel movement frequency didn’t differ significantly between groups, symptoms such as incomplete evacuation and abdominal pain before defecation did improve in the probiotic group. This finding contrasts with many previous probiotic trials where increased stool frequency was often the primary indicator of success. For instance, Majeed et al. reported that *B. coagulans* Unique IS2 boosted both stool frequency and consistency in individuals with functional constipation [22]. On the other hand, Wang et al. found that while *B. bifidum* CCFM16 increased stool frequency, it didn’t lead to improvements in quality-of-life scores like PAC-QOL or PAC-SYM [23]. Other trials, such as those by Chmielewska et al. and studies involving *L. casei* Shirota, also showed increased stool frequency without meaningful changes in stool quality or abdominal discomfort [24, 25]. These findings highlight a critical nuance: changes in stool frequency alone may not be enough to improve the lived experience of bowel issues.

The multi-strain probiotic used in this study likely owes its broader impact to the diverse and complementary roles of its component strains: *Clostridium butyricum* IDCC 1301, *Weizmannia coagulans* IDCC 1201, and *Bacillus subtilis* IDCC 1101. *C. butyricum* is known for producing butyrate, a short-chain fatty acid that nourishes intestinal epithelial cells, supports mucosal integrity, and helps regulate gut permeability [26]. It also lowers intestinal pH, discouraging the growth of harmful bacteria [27]. In our study, reductions in abdominal pain prior to defecation (p = 0.0090) may be tied to these butyrate-mediated effects, which have been shown to support mucosal health and dampen inflammation [28, 29]. Some evidence even suggests that *C. butyricum* can influence the gut–brain axis, potentially alleviating visceral pain through neural modulation [30–32].

Meanwhile, *W. coagulans* produces lactic acid, which stimulates intestinal peristalsis and maintains an acidic environment that deters pathogens [33, 34]. It also appears to play a role in modulating immune responses and reducing gut inflammation [33]. In our trial, participants taking the probiotic blend reported fewer straining episodes during defecation (p = 0.0458), which may be linked to enhanced motility. The sense of incomplete evacuation also decreased significantly (p = 0.0374), likely due to improved bowel emptying. Previous research supports these findings, showing similar benefits from *W. coagulans* supplementation [35].

Lastly, *B. subtilis* contributes by producing digestive enzymes that aid nutrient breakdown and encourage the growth of beneficial gut bacteria such as *Lactobacillus* and *Bifidobacterium* [36]. It also helps create a healthier gut environment by inhibiting harmful microbes [37]. Participants in the probiotic group were more likely to report normal stool forms (Bristol types 3–4; p = 0.0176), possibly due to improved digestion and stool formation supported by these enzymes [38].

Altogether, while many single-strain probiotics tend to focus on stool frequency, this multi- strain blend appears to offer more comprehensive relief—addressing symptom severity and quality of life. From a patient perspective, reducing discomfort, straining, or feelings of incomplete evacuation often matters more than just increasing the number of bowel movements. While some effects could be attributed to placebo, the clear between-group differences in symptom relief suggest a genuine therapeutic benefit.

No serious adverse events were reported during the trial, indicating good tolerability and safety within the study population. Still, this study has limitations. The relatively small sample size (n = 78) limits the ability to generalize the findings. Also, the four-week intervention period captures only short-term effects, leaving questions about long-term efficacy unanswered. We also didn’t examine gut microbiota composition or metabolite changes, which could offer deeper insight into how these probiotics exert their effects.

Future studies should involve larger and more diverse populations, extend the duration of intervention, and incorporate microbiome and metabolome analyses. These steps will be essential to fully understand the mechanisms and long-term benefits of this probiotic blend.

## Conclusions

The findings of this study suggest that 4-week spore-forming probiotic blend supplementation may improve digestive health in healthy adults with mild gastrointestinal discomfort by reducing defecation-related discomfort and enhancing stool consistency. Significant improvements were observed in abdominal pain before bowel movements, irritant bowel movements, and the sensation of incomplete evacuation, while stool form shifted toward a more optimal consistency. Furthermore, a significant increase in physical functioning scores was observed, suggesting a potential link between digestive health and overall well-being.

While these results suggest that probiotic blend may support gut health, further research is needed to confirm these findings and elucidate the underlying mechanisms. A longer intervention period and comprehensive microbiome analysis would provide additional insights into its effects, enabling a more robust evaluation of its role in digestive health management.

## Data Availability

All relevant data are within the manuscript and its Supporting Information files

## Acknowledgments

We thank all volunteers for their participation and commitment to this clinical study. We also acknowledge the valuable support of the Clinical Trial Center of Kyung Hee University Korean Medicine Hospital for subject management and clinical coordination.

## Author Contributions

**Data curation:** Hyung Gyu Park, Han Bin Lee, Haeseong Park

**Formal analysis:** Hyung Gyu Park, Han Bin Lee, Haeseong Park

**Investigation:** Minji Kang, Minkyung Bok, Yeongtaek Hwang, Sungho Maeng

**Methodology:** Minji Kang, Minkyung Bok, Yeongtaek Hwang, Sungho Maeng

**Project administration:** Kyuho Jeong

**Funding acquisition:** Kyuho Jeong

**Supervision:** Jin Seok Moon, Hyunjung Lim

**Writing – original draft:** Hyung Gyu Park, Han Bin Lee, Haeseong Park

**Writing – review & editing:** Jin Seok Moon, Hyunjung Lim

**Data Availability Statement:** All relevant data are within the manuscript and its Supporting Information files.

## Supporting information

**S1 Table. Compositions of the Probiotic blend and Placebo Products. S2 Table. Daily Intake of Energy and Nutrients.**

**S3 Table. Changes in Physical Activity. S4 Table. Changes in Bowel Function.**

